# “Transcriptomic Profiling Unveils Novel Therapeutic Options for Drug-Resistant Temporal Lobe Epilepsy”

**DOI:** 10.1101/2024.06.26.24309519

**Authors:** Patricia Sánchez-Jiménez, Lola Alonso-Guirado, Laura Cerrada-Gálvez, Marcos Elizalde-Horcada, Inmaculada Granero-Cremades, Paolo Maietta, Antonio Gómez-Martín, Francisco Abad-Santos, Cristina Virginia Torres-Díaz, Maria de Toledo, Natalia Frade-Porto, Patricia Gonzalez-Tarno, Paloma Pulido, María C. Ovejero-Benito

## Abstract

**Background:** Epilepsy drug treatments fail in 25-30% of patients, who then develop drug resistance. Temporal lobe epilepsy is the most prevalent subtype associated with drug resistance. Classical drug discovery is a long and extremely costly process with a high rate of failure in clinical trials. Drug repurposing is a more cost- and time-effective strategy. Hence, the main objective of this study is to propose drug candidates for the treatment of drug-resistant temporal lobe epilepsy (DR-TLE) through drug repurposing based on transcriptomic profiling.

**Methods:** Total RNA-sequencing (RNA-Seq) was performed on 45 formalin-fixed paraffin-embedded (FFPE) hippocampi of DR-TLE patients and 36 FFPE hippocampi of post-mortem biobank donors. RNA-Seq was carried out in an Illumina NovaSeq 6000 platform in 100bp paired-end. Drug repurposing based on transcriptomic analysis top candidates was performed against these databases: Pandrugs2, PharmOmics, DGIdb, ToppGene, L1000CDS^2^ and Connectivity Map.

**Results:** We found 887 genes differentially expressed between DR-TLE patients and post-mortem controls. We observed 74 potential drug candidates in at least two independent databases. Of these, we selected only the 11 which can cross the blood-brain barrier: cobimetinib, panobinostat, melphalan, rucaparib, alectinib, ponatinib, danazol, carboplatin, vandetanib, erlotinib, and gefitinib. After analyzing their safety and efficacy profile based on previous publications, we provide a list of the top 5 candidates.

**Conclusions:** We therefore propose erlotinib, danazol, rucaparib, ponatinib, and panobinostat as potential therapies for DR-TLE based on differential RNA-Seq profiling.

## INTRODUCTION

Epilepsy is a chronic neurological disease, often difficult to detect, frequently of unknown etiology and challenging to control. The number of approved antiseizure drugs (ASDs) has increased considerably over the last few years ^1,2^. However, 25-33% of epileptic patients do not respond adequately to these medications ^1^, and they develop drug-resistant epilepsy (DRE). Frequently, neurologists observe DRE appearing in focal epilepsies, especially in those associated with temporal lobe epilepsy (TLE), the most common subtype of epilepsy ^3^. Nowadays, the etiology of DRE is still unknown, but it has been found that drug resistance often occurs nine years after starting pharmacological treatment ^4^ although the cause of this latency is still unknown. Several hypotheses try to explain DRE, such as genetic variations, disease-related and drug-related mechanisms ^5^. It has been hypothesized that environmental factors and epigenetic mechanisms are involved in the development of drug resistance ^6–11^. Epigenetic modifications can modulate gene expression, and different studies have explored the connection between gene expression and drug resistance ^12–17^. There are several therapeutic alternatives for DRE patients, such as the highly effective neurosurgical resection of the epileptogenic zone ^18^. However, this is only applicable to focal epilepsies such as TLE ^19^. In addition, there is an unmet need for disease-modifying ASDs that reduce the severity or delay the onset of seizures ^20^.

Discovery of new ASDs is a long and incredibly costly process, with a high failure rate in clinical trials due to inefficacy ^21^. The failure of new drugs for epilepsy in clinical trials may be partly explained by the fact that ASDs are tested in preclinical models such as maximal electroshock seizure (MES) and s.c. pentylenetetrazole (PTZ). These could be considered models of seizure rather than models of epilepsy ^22^. Drug discovery is implementing computation models and new technologies to accelerate the process ^23–26^. However, the time period from candidate drug to approved drug is still long and with a low success rate ^27^. In recent years drug repurposing strategies have been used to identify new indications for approved drugs ^28^. Drug repurposing accelerates the approval of these new treatments by 3 to 12 years and reduces costs and risk by 50%. The approval rate is higher in repurposed drugs (30%) than in new compounds (10%) ^2^. Drug repurposing is based on the fact that drugs may have multiple targets, while different diseases may have in common the same genetic factors, molecular pathways, and/or clinical manifestations. Therefore, a drug can potentially modulate these common factors and may benefit different diseases ^29^. Drug repurposing has been successfully used for diverse diseases such as cancer or COVID-19 ^30–32^.

Systematic analysis of classical drug discovery pipelines showed that drugs developed with supporting human genetic data had a double probability of being approved ^21,33^. This approach has not been deeply explored in the epilepsy field, since, to our knowledge, there is only one study of drug repurposing based on transcriptomics in epilepsy ^34^.

The main objective of this study is the identification of approved drug candidates that can modulate factors and/or molecular pathways involved in drug-resistant temporal lobe epilepsy (DR-TLE) based on the differential gene expression data.

## MATERIALS AND METHOD

### Study population

A total of 46 hippocampal formalin-fixed paraffin-embedded (FFPE) samples from DR-TLE patients subjected to neurosurgery in the previous ten years were analyzed. All the recruited patients agreed with the protocol and signed the informed consent approved by the Independent Clinical Research Ethics Committee of Hospital Universitario de La Princesa. The study was carried out according to the revised Declaration of Helsinki and STROBE guidelines.

Thirty-five hippocampal FFPE samples were provided as control samples from Spanish biobanks: “Biobanco en Red de la Región de Murcia” (BIOBANC-MUR), “Biobanco del Hospital Universitario Puerta de Hierro” (HUPHM), “Biobanco del Instituto de Investigaciones Biomédicas August Pi i Sunyer” (IDIBAPS), and “Biobanco de Navarrabiomed” (Navarrabiomed). Moreover, they were processed following standard operating procedures with the approval of their Ethics and Scientific Committees. None of them suffered any psychiatric, neurological, or neurodegenerative diseases, and all were analyzed by anatomopathologist to discard pathological features. Controls were of similar age to the patients.

### Sample processing and RNA isolation

A total of 81 hippocampal FFPE blocks were cut on a 15μm scroll. RNA was isolated with the truXTRAC® FFPE total NA kit - Column (Covaris, USA), according to the manufacturer’s instructions. Then, DNA-free RNA was obtained with the RNase-Free DNase Set (Qiagen, Germany) and the RNeasy® MinElute® Cleanup kit (Qiagen, Germany). Quantification and quality control were performed before and after those steps using the 4200 TapeStation system (Agilent, USA) and the Qubit 2.0 Fluorometer (Thermo Fisher Scientific, USA).

### Library Preparation and Sequencing

Libraries were prepared using the SMARTer® Stranded Total RNA-Seq kit v2-Pico Input Mammalian (Takara Bio, Japan) following the manufacturer’s protocol to process FFPE samples. Libraries were sequenced to achieve an average output of 50M paired-end reads (100bp pair-end) on the Illumina NovaSeq^TM^ 6000 platform. Read quality was assessed by fastp ^35^. Raw reads were aligned against GRCh37/hg19 reference genome with HISAT2 ^36^ and StringTie ^37^ to obtain a counts matrix according to the protocol described by ^38^.

### Differential Expression Analysis using DESeq2

We performed a differential expression analysis of genes using the DESeq2 package (version 1.34.0) in the R software (R version 4.2.2). Raw gene counts matrix was pre-processed before the analysis. In the quality control checks performed, samples with an alignment rate lower than 5% were discarded (13 controls and 1 patient). The sample size was reduced from 81 (35 controls and 46 patients) to 67 subjects (22 controls and 45 patients) to satisfy our integrity and accuracy quality thresholds (Figure 1).

**Figure 1:**
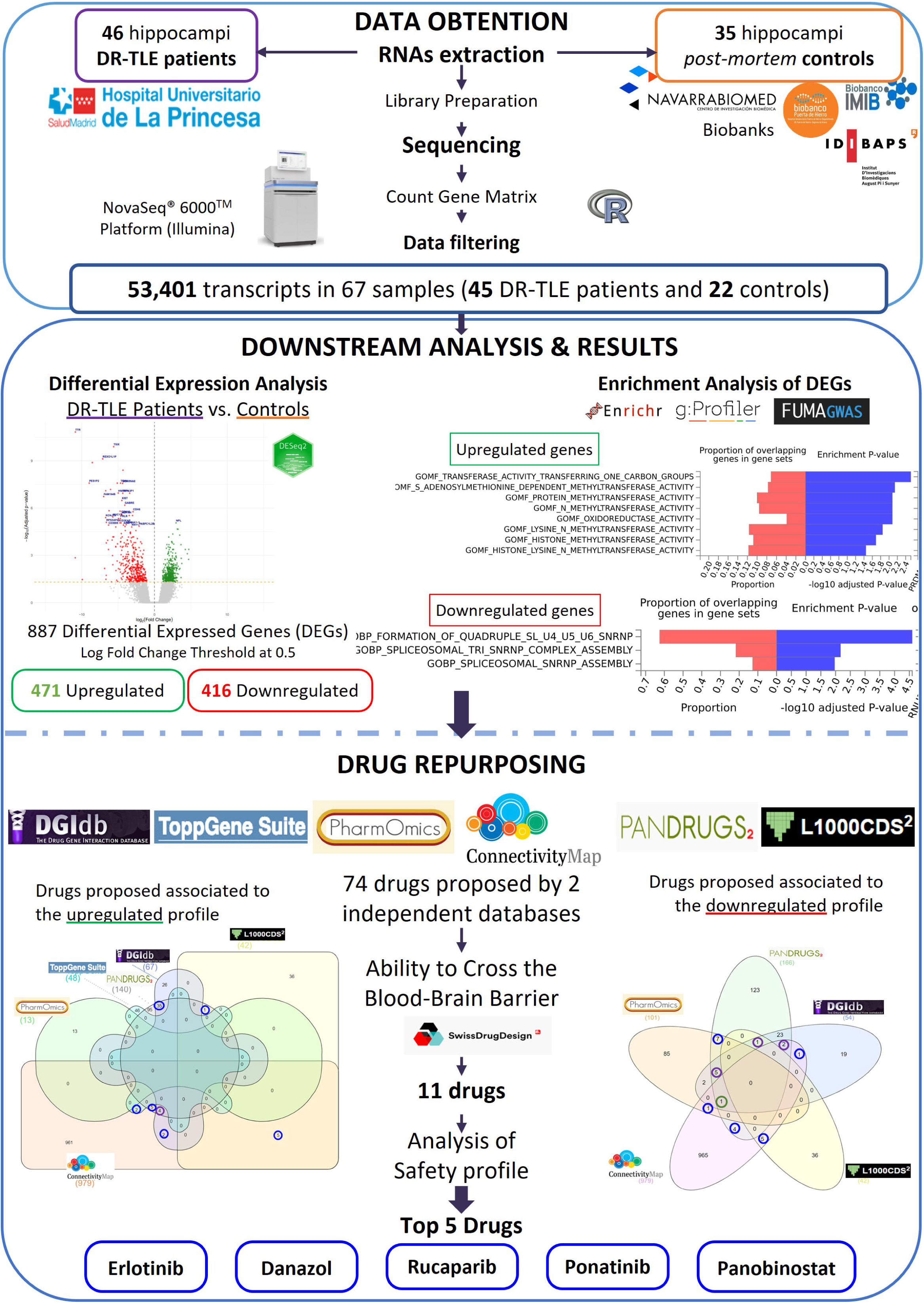
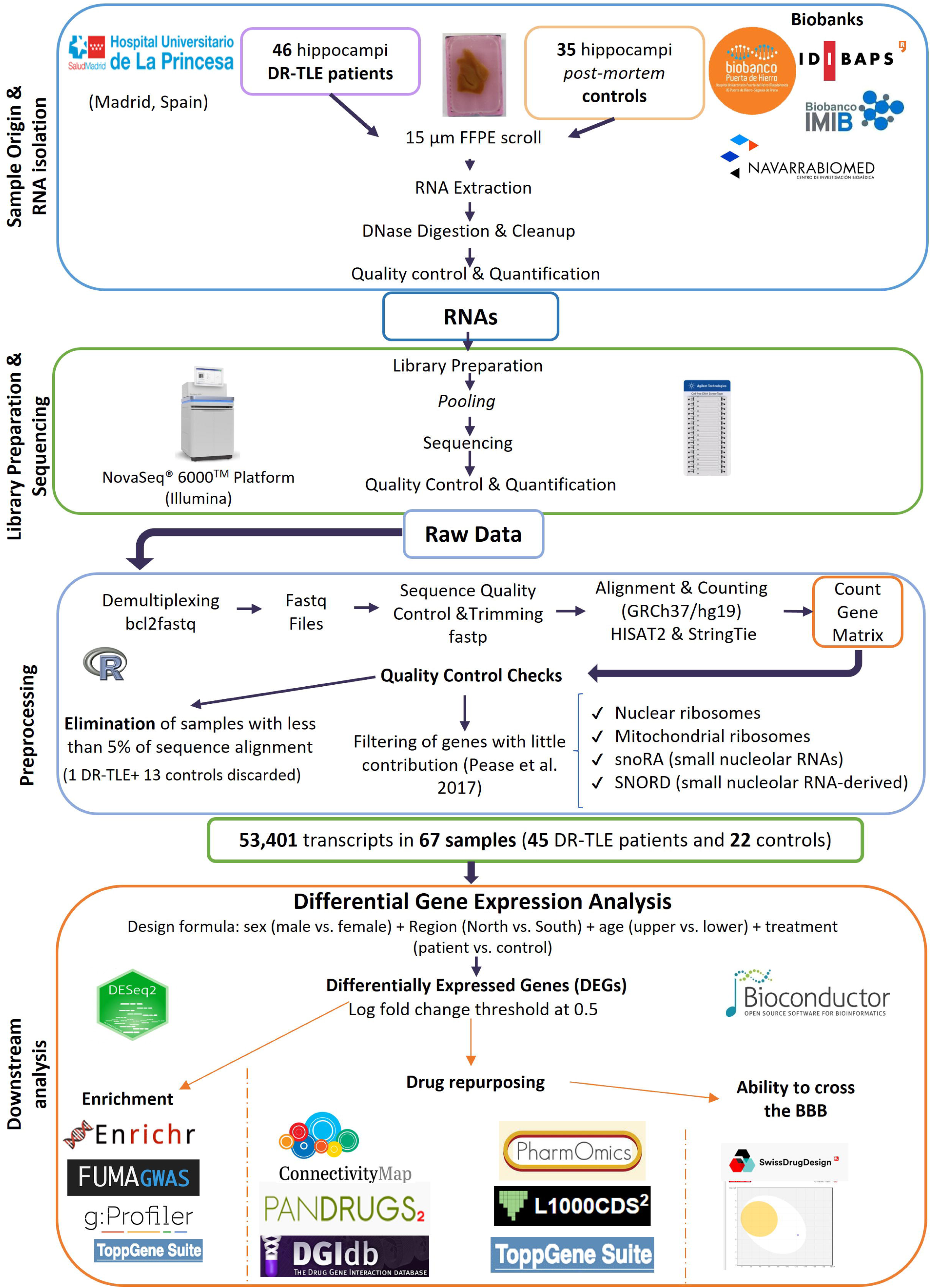
Workflow of the experiments and analysis carried out. Abbreviations: BBB: blood-brain barrier; CMap: Connectivity Map; DGIdb: Drug Gene Interaction Database; DR-TLE: drug-resistant-temporal lobe epilepsy.

We loaded the complete phenotypes dataset from ENSEMBL 75, and we filtered out nuclear and mitochondrial RNA-ribosomes, snoRA (small nucleolar RNAs), and SNORD (small nucleolar RNA-derived) genes ^39^ from the gene raw counts dataframe. The raw count data was then normalized using the DESeq2 normalization methods. As “metadata” information about the samples, we had alignment rate, cohort information, sex or batch. To control the potential effect of geographical regions on the study outcomes, a new variable called “region” was included in the contrast parameters. Thus, the contrast parameters used for the DESeq2 analysis were the variables “sex”, “region”, “age”, and “treatment”. Then, we obtained the list of differentially expressed genes (DEGs) with an absolute lfcThreshold=0.5 and a FDR-adjusted p-value lower than 0.05.

### Enrichment analysis

Signaling pathways and biological functions associated with DR-TLE were analyzed using the following tools: Enrichr ^40^, FUMA GWAS ^41^, g:Profiler ^42^ and ToppGene ^43^. We analyzed separately for the upregulated and downregulated genes in patients with respect to controls (Figure 1). Only significant results with a FDR-adjusted p-value <0.05 were considered.

Protein network analysis was performed by STRING ^44^ and Cytoscape software version 3.10.1. ^45^. Markov Clustering Algorithm was applied to determine groups of nodes within the network using a 0.4 cutoff and 4 points of granularity.

### Drug repurposing

Drug repurposing was performed using the following tools: Pandrugs2 ^46^; PharmOmics ^47^; Drug Gene Interaction Database (DGIdb) ^48^; ToppGene Suite ^43^; L1000CDS^2^ ^49^; Connectivity Map (CMap) ^50^ (Figure 1). All these algorithms score the association between genes and drug targets.

Some drug candidate lists were filtered more restrictively to select the top candidates. In Pandrugs2, just the best candidates for therapies were selected. The functions of the proteins encoded by downregulated genes in DR-TLE patients with respect to controls are decreased in these patients. Therefore, we filtered the list of drugs proposed by DGIdb to modulate downregulated genes, and discarded drugs classified as blockers, inhibitors, or antagonists. In Connectivity Map (CMap), the drug candidates with negative Raw_cs and a base 10 logarithm of FDR (fdr_q_nlog10) higher than 15.35, were selected. CMap allows a maximum input of only 150 upregulated and 150 downregulated DEGs. To perform this analysis, we selected those genes with the lowest and highest log2fc values and that were included in the CMap database.

Venn Diagram plots were performed with the tool Interactive Venn (https://www.interactivenn.net/).

To increase the reliability of our results, the webtool SwissADME ^51^ was employed to predict whether potential drugs could cross blood-brain barrier BBB and access the target tissue. This tool also analyzes whether drugs are substrates of Glycoprotein-P that reduces the amount of drugs that reach the brain ^5,52^, and the ability of these drugs to inhibit CYP450 superfamily enzymes.

Pan-Assay Interference compounds (PAINS) are promiscuous molecules that form unspecific interactions with many different proteins. For this reason, PAINS could give rise to false positives in drug repurposing studies ^53^. We filtered out the PAINS identified with the webtool Hit Dexter 3 ^53^.

## RESULTS

### Study population

Initially, 81 samples were recruited (46 DR-TLE patients and 35 controls). Our final cohort after having discarded the samples that did not reach quality standards included 67 subjects: 45 DR-TLE patients, with a mean age of 46.20 ± 10.36 years old, and 22 post-mortem controls, with a mean age of 49.09 ± 14.80 years old (Table 1). The percentage of men in the patient group was 46%, while in the control group was 74% (Table 1), representing a significant difference (p=0.045). This imbalance increased because some samples were removed from the initial cohort after quality control checks. The main clinical and demographic characteristics of patients and controls are described in Table 1.

**Table 1:** Summary of the clinical and demographic characteristics of the study population and the controls.

|  |  | DR-TLE | Controls |
| --- | --- | --- | --- |
| Number |  | 46 | 35 |
| Men (%) |  | 24 (46%) | 26 (74%) |
| Age at surgery/Age at death (years) |  | 46.09 ± 10.27 | 52.94 ± 14.14 |
| Age of onset (years) |  | 14.63 ± 13.86 | NA |
| Febrile seizures in childhood |  | 16 (35%) | NA |
| Number of seizures/month |  | 13.15 ± 22.38 | NA |
| Aura (%) |  | 31 (67%) | NA |
| Number of anti-seizure drugs |  | 5.96 ± 3.39 | NA |
| Cranioencephalic traumatism (%) |  | 5 (11%) | NA |
| Sensory, gait, or developmental disturbances (%) |  | 10 (22%) | NA |
| Ever smoker (%) |  | 18 (39%) | NA |
| Alcohol consumption (%) |  | 13 (28%) | NA |
| Seizure types | Focal (%) | 32 (70%) | NA |
|  | Generalized (%) | 2 (4%) | NA |
|  | Combined generalized and focal (%) | 10 (22%) | NA |
| Seizure etiology | Structural (%) | 3 (8%) | NA |
|  | Infectious (%) | 13 (33%) | NA |
|  | Unknown (%) | 24 (60%) | NA |
| Seizures signs | Absence (%) | 24 (52%) | NA |
|  | Convulsion (%) | 10 (22%) | NA |
|  | Both (%) | 12 (26%) | NA |
| Right dominant hand (%) |  | 33 (72%) | NA |
| Surgically intervened hemisphere | Right (%) | 21 (46%) | NA |
|  | Left (%) | 25 (54%) | NA |
| Engel (12 months) | R (%) | 43 (93%) | NA |
|  | NR (%) | 3 (7%) | NA |
| Severe adverse effects (12 months) (%) |  | 4 (9%) | NA |
| Engel (24 months) | R (%) | 43 (93%) | NA |
|  | NR (%) | 3 (7%) | NA |
| Severe adverse effects (24 months) (%) |  | 2 (4%) | NA |

Data are shown as mean ± SD or number and percentage. Abbreviations: DR-TLE: drug-resistant temporal lobe epilepsy; NA: not applicable; NR: non responder to neurosurgery (Engel III or IV); R: responder to neurosurgery (Engel I or II).

### Differentially expressed genes associated with DR-TLE

We observed 887 DEGs between DR-TLE patients and *post-mortem* controls: 471 upregulated and 416 downregulated in patients with respect to controls (Figure 2A, Supplementary Table 1). The top 10 upregulated and downregulated genes in DR-TLE patients with respect to controls are shown in Table 2.

**Figure 2.**
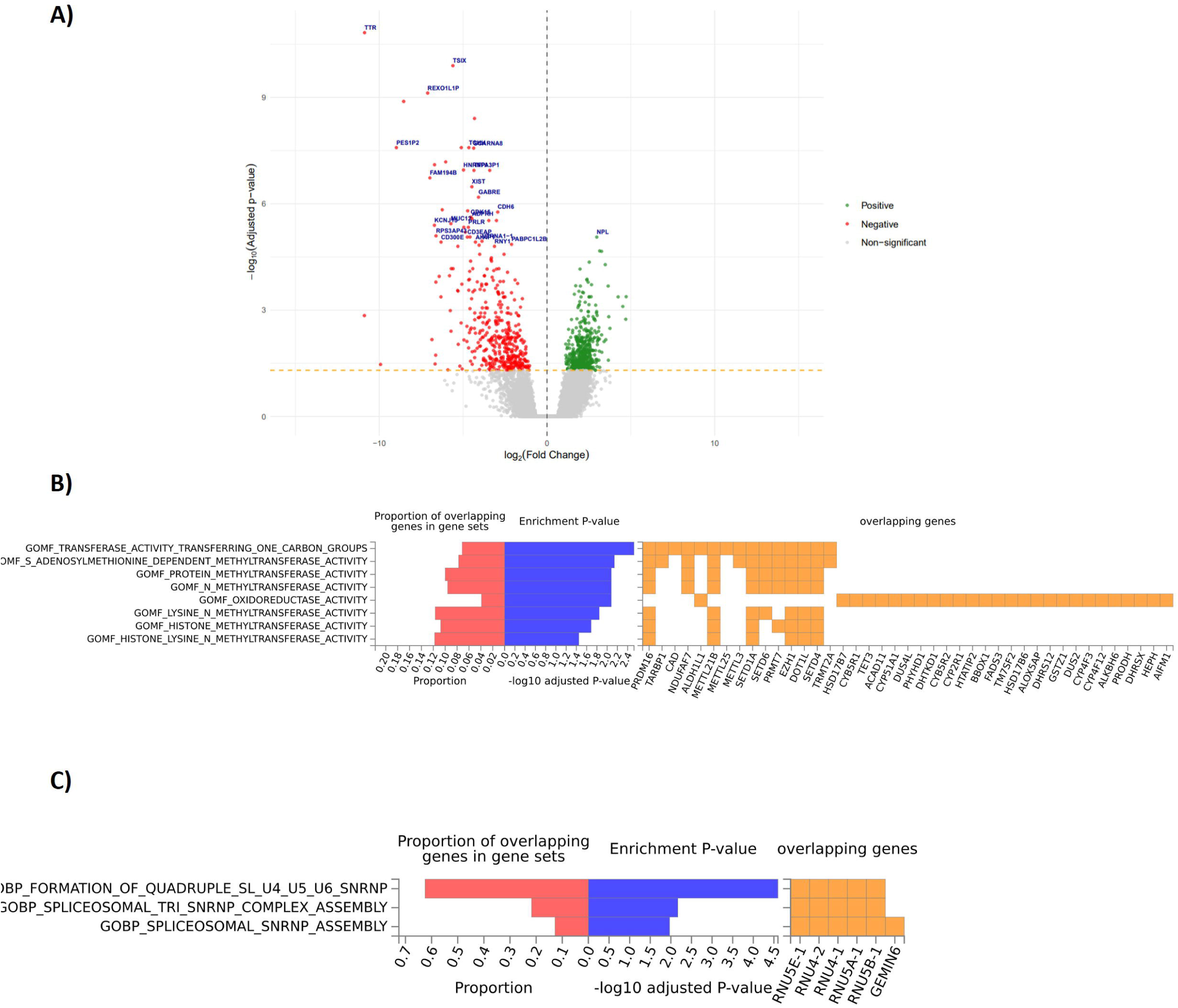
**A)** Volcano plots of the DEGs associated with DR-TLE in the hippocampus. The logarithm of the association p-value for each DEGs is displayed along the Y-axis while the logarithm of fold change is shown along the X-axis. Green and red dots show DEGs over-expressed and under-expressed in patients with respect to controls, respectively. Blue dots show DEGs that do not have a significant association with DR-TLE. Only the names of the 40 DEGs with the lowest adjusted p-value are displayed. B) Enrichment analysis performed with FUMA GWAS analyzing functions of the DEGs in the hippocampus according to the GO molecular function and GO biological processes libraries. Red bars represent the proportion of overlapping genes in the gene set. Blue bars show the enrichment p-value, represented as the logarithm of the FDR adjusted p-value. Orange squares show the genes involved in every enrichment term. Abbreviation: FDR, false discovery rate. **B)** Enrichment analysis of the DEGs upregulated in patients with respect to controls. **C)** Enrichment analysis of the DEGs downregulated in patients with respect to controls. Red bars represent the proportion of overlapping genes in gene set. Blue bars show the enrichment p-value, represented as the negative logarithm of the FDR adjusted p-value. Yellow squares show the genes involved in every enrichment term. Abbreviation: DEGs: Differentially Expressed Genes; DR-TLE: drug-resistant temporal lobe epilepsy; FDR: false discovery rate.

**Table 2:**
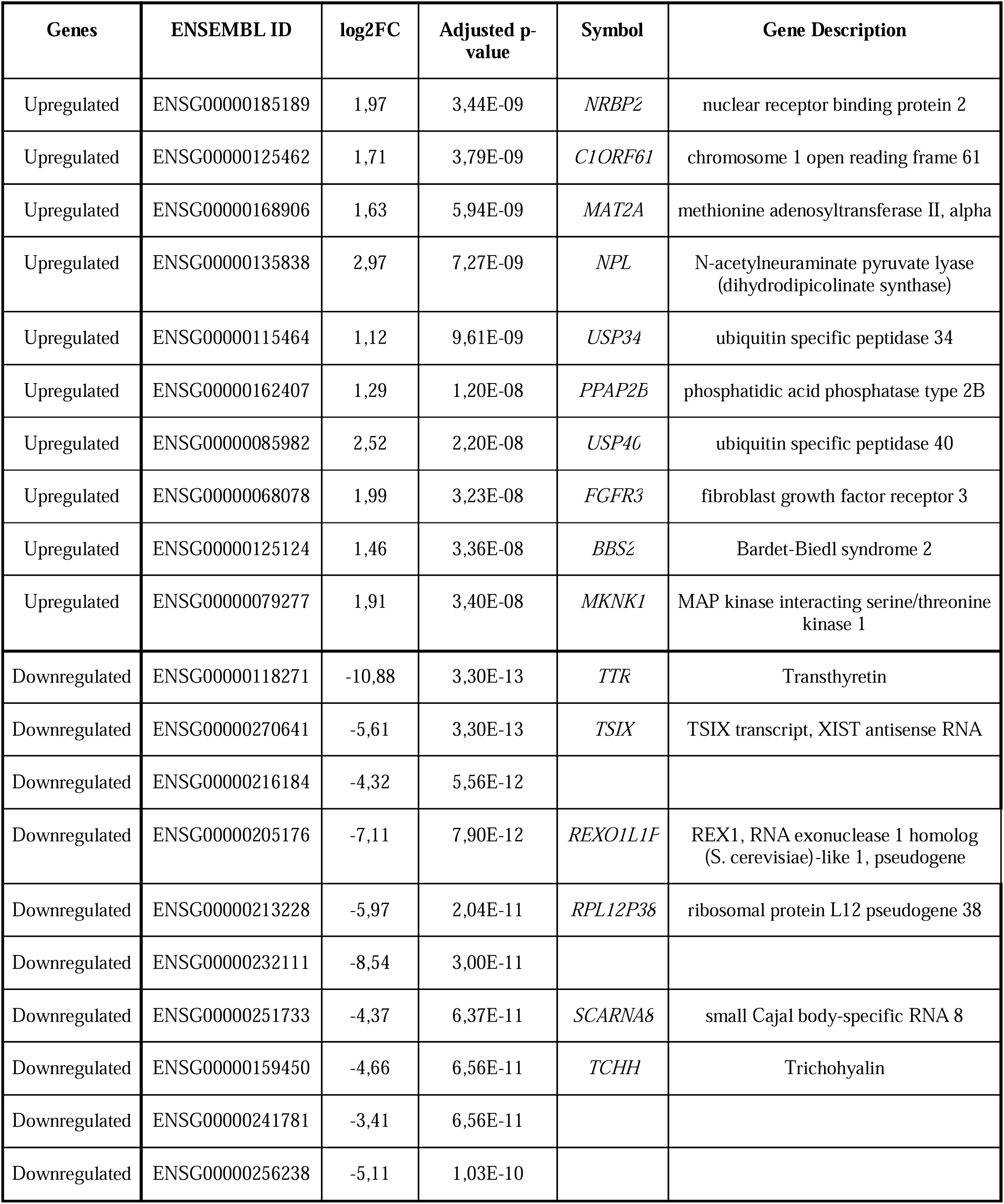
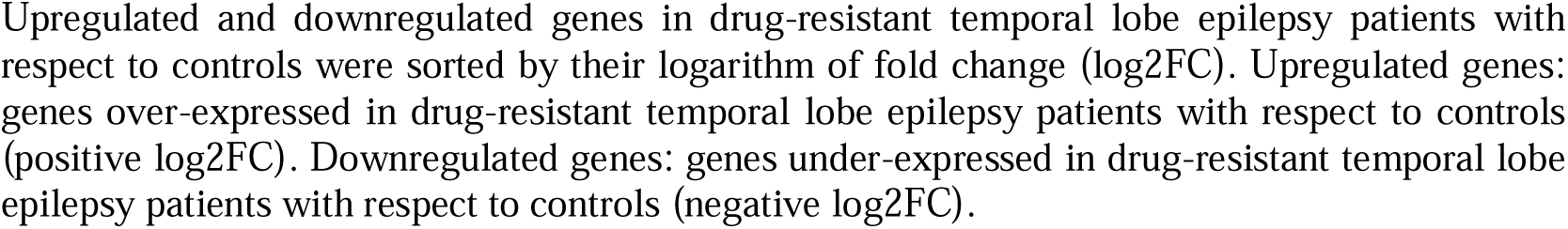
Top 10 upregulated and downregulated genes in drug-resistant temporal lobe epilepsy patients with respect to controls.

### Enrichment analysis

To determine the biological functions and the signaling pathways where the DEGs associated with DR-TLE were involved, we performed an enrichment study. Genes upregulated in DR-TLE were enriched in methyltransferase activity, oxidoreductase activity, catabolic processes, lipid metabolism and immune system activation (Figure 2B, Supplementary Figure 1B and 1C). In contrast, downregulated genes were enriched in alternative splicing, calcium ion binding, and Cajal bodies (Figure 2C, Supplementary Figure 1A).

Furthermore, we performed a clustering analysis with Cytoscape of the proteins encoded by DEGs associated with DR-TLE. This software grouped proteins in clusters of 5 or more, related to specific functions (Figure 3, Supplementary Table 2). Upregulated genes were clustered in 8 groups (Supplementary Table 2A), with the four largest involved in functions such histone modifications (Figure 3A-1), metabolic pathways (Figure 3A-2), inflammatory response (Figure 3A-3), and cysteine and methionine metabolism (Figure 3A-4). Downregulated genes were grouped in 5 clusters (Supplementary Table 2B). The four biggest clusters were implicated in functions such as calcium signaling (Figure 3B-1), cytoplasmic ribonucleoprotein granule (Figure 3B-2), PI3K-Akt signaling pathway (Figure 3B-3), and eosinophil percentage of leukocytes (Figure 3B-4, Supplementary Table 2B).

**Figure 3.**
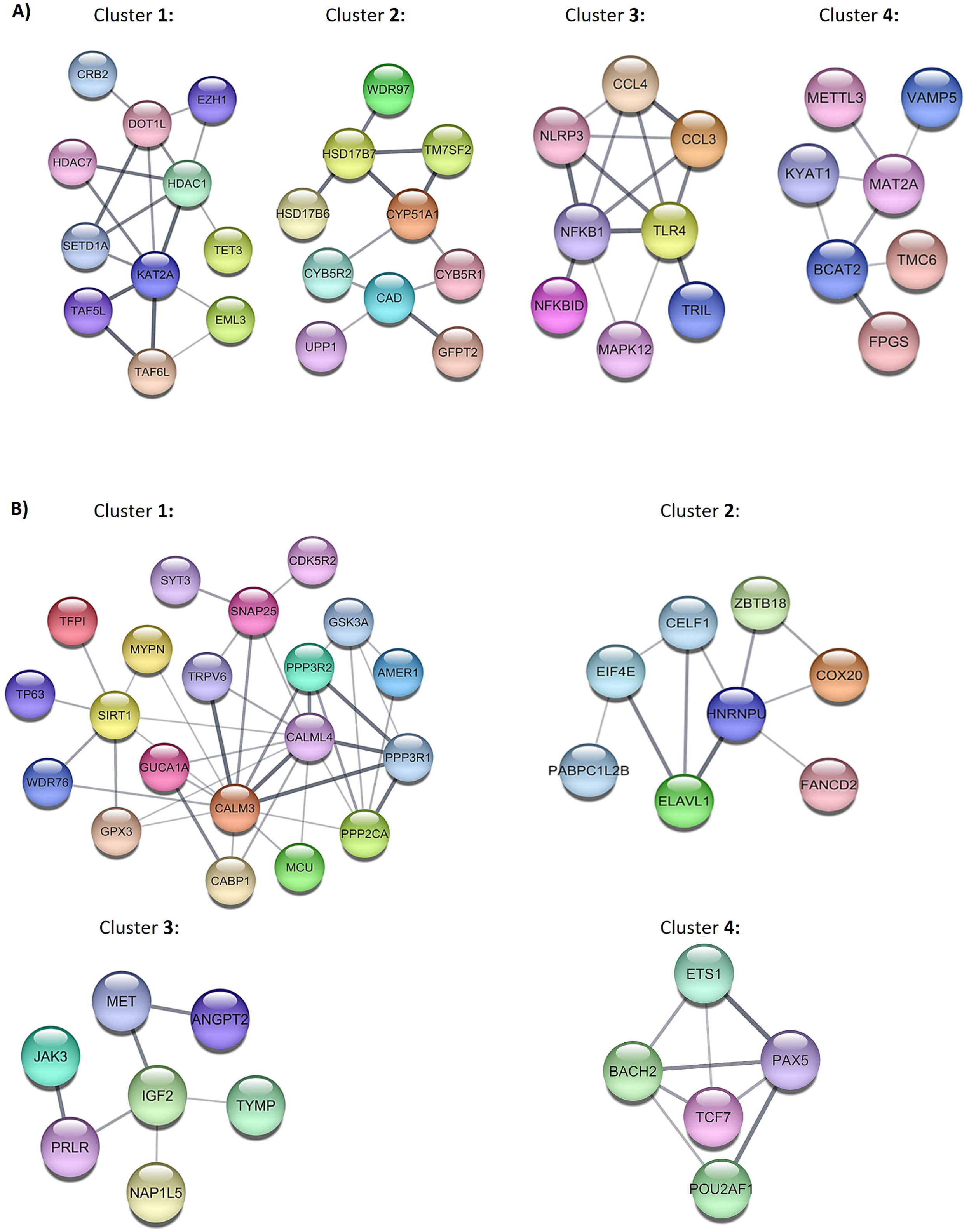
Cytoscape network of proteins expressed by DEGs associated with DR-TLE. A) Top 4 largest clusters of the proteins encoded by the genes upregulated in DR-TLE patients with respect to controls. B) Top 4 largest clusters of the proteins encoded by the downregulated in DR-TLE patients with respect to controls. Abbreviation: DEGs: Differentially Expressed Genes; DR-TLE: drug-resistant temporal lobe epilepsy.

### Drug repurposing

We found 42 potential drug candidates in L1000CDS^2^, and 979 in CMap, associated with both upregulated and downregulated genes (Supplementary Figure 2A, Supplementary Table 3A). In the case of **upregulated** DEGs in DR-TLE patients with respect to controls, we observed 140 potential drug candidates in the Pandrugs2 database, 13 in PharmOmics, 67 in DGIdb and 48 in ToppGene (Supplementary Table 3A). Then, we analyzed the 54 candidates identified by at least two different databases (Supplementary Table 3B). We observed 35 potential drug candidates in Pandrugs2 and DGIdb; 1 in both Pandrugs2 and L1000CDS^2^; 5 in Pandrugs2 and CMap; 5 in L1000CDS^2^ and CMap; 2 in both DGIdb and CMap; 2 in both ToppGene and CMap and 4 in Pandrugs, DGIdb and CMap (Supplementary Figure 2B, Supplementary Table 3B). Only 9 of these potential drug candidates cross the BBB (Supplementary Table 3C).

After that, we searched for potential drugs that could modulate **downregulated** genes in DR-TLE patients with respect to controls. We obtained 166 potential drug candidates in Pandrugs2; 101 in PharmOmics; and 54 in DGIdb (Supplementary Figure 2B, Supplementary Table 3D). ToppGene did not propose any drug candidate for downregulated genes. Again, we focused on the 52 candidates proposed by at least 2 different databases. We obtained 7 potential drug candidates in PharmOmics and Pandrugs2; 2 in both PharmOmics and DGIdb; 1 in PharmOmics and CMap; 23 in Pandrugs2 and DGIdb; 4 in Pandrugs2 and CMap; 5 in L1000CDS^2^ and CMap; and 1 in DGIdb and CMap (Supplementary Figure 2B, Supplementary Table 3E). We also found 8 potential drug candidates proposed by 3 databases: 5 drugs in both PharmOmics, Pandrugs2 and DGIdb; 1 in Pandrugs2, DGIdb, and L1000CDS^2^; and 2 in Pandrugs2, DGIdb, and CMap (Supplementary Figure 2B, Supplementary Table 3E). We discovered 1 potential drug candidate in common in 4 databases: docetaxel was proposed by PharmOmics, Pandrugs2, DGIdb, and CMap (Supplementary Figure B, Supplementary Table 3E). From all the mentioned drug candidates, we selected only the 9 drugs that cross the BBB (Supplementary Table 3F).

We then explored the pharmacokinetics, pharmacodynamics, and toxicology properties of the drug candidates. To select the best candidate drugs for treating DR-TLE, we focused on those which can reach the brain. We studied the therapeutic indication, target, and mechanism of action of these drugs (Supplementary Table 4). As the repurposed drugs may be used in combination with other ASDs, we analyzed the ability of the candidate drugs to inhibit the main enzymes of cytochrome P450 superfamily (Supplementary Table 4). Finally, we described the most common adverse drug reactions (ADRs) that appear in one in ten patients (Very Common ADRs) (Supplementary Table 4). Based on their pharmacological and toxicological profile, we obtained a list of the top 5 candidates for treating DR-TLE: erlotinib, danazol, rucaparib, ponatinib, and panobinostat (Table 3).

**Table 3:**
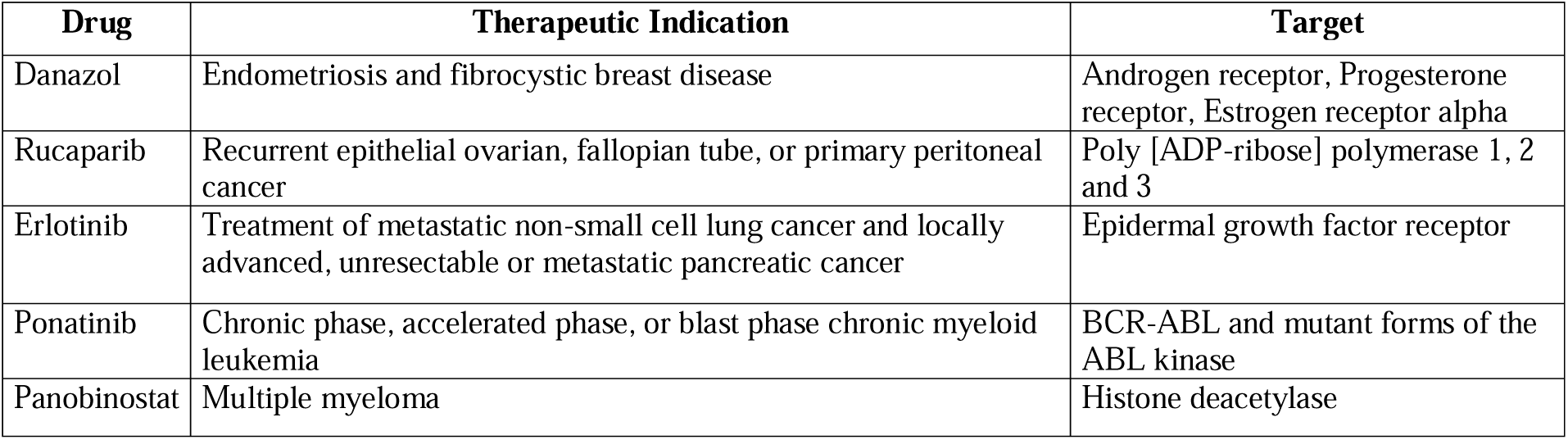
List of the top five drug candidates proposed for drug-resistant temporal lobe epilepsy treatment.

## DISCUSSION

This study applied next-generation sequencing to the epileptogenic zone of DR-TLE patients in order to guide the selection of drug candidates, already approved to treat other conditions, for the treatment of this disease. We observed 471 upregulated and 416 downregulated genes in DR-TLE patients compared to *post-mortem* non-epilepsy controls. We observed that genes upregulated in DR-TLE were involved in different processes such as histone modifications, methyltransferase activity, metabolic pathways, inflammatory response, alternative splicing, lipid oxidation or cysteine metabolism. Most of these processes have been previously associated with DRE ^6,54–56^. In fact, the over-expression of histone modification and methyltransferases may support the epigenetic hypothesis of DRE ^6,57–59^. There is wide evidence that inflammatory response is deeply involved in DRE ^5,7,8,60–62^. Moreover, mutations on splicing sites are associated with myoclonic epilepsy ^56^ or epileptic encephalopathies ^63^. In addition, lipid metabolism and fatty acid oxidation were overexpressed in brain samples of DRE patients ^55,64^. Cysteine and methionine metabolism were upregulated in DR-TLE patients. Surprisingly, cysteine levels are increased in blood samples of epilepsy patients ^54^. On the other hand, genes downregulated in DR-TLE patients play roles related to DRE such as calcium signaling ^5^, senescence or PI3K signaling ^65^.

There are previous transcriptomic analysis of DRE and DR-TLE ^12–15,17,61,66–68^. Some of these took a different approach from this study, focusing on specific features such as hippocampal sclerosis ^14,17^, febrile seizure ^68^, or electric activity ^69^. Other studies analyzed different brain regions such as the neocortex ^15,69^. Discrepancies in the number of DEGs found in these studies could be explained partially by diverse factors, such as patients’ ethnicity ^12^, the technique used ^69^, or the conservation method (paraffin-embedded and fixed versus fresh tissue) ^14,15,17^. We used FFPE tissue based on the availability of these samples in the anatomical pathology unit of the hospital and biobanks. However, paraffin may act as an intercalant in nucleic acids, modifying nucleotides and affecting RNA integrity and stability ^70^. Thus, we had to discard 14 samples (13 controls and 1 DRE patient) because their alignment rate was lower than 5%. Even after this patient exclusion, we had a sample of 46 patients, a higher number than in previous studies ^12,14,61^.

Drug repurposing based on transcriptomics is a powerful, time- and cost-effective strategy ^30–32^. In epilepsy, the principal strategy to discover new candidates for drug therapy was genome-wide association studies ^30,71^. In this study, we used the transcriptomic strategy for drug repurposing and employed six repurposing databases. We selected drugs proposed by at least two independent databases to increase the robustness of the results ^32^. Epilepsy treatment is challenging because BBB limits the number of drugs that reach the brain ^72^. However, we found 11 drug candidates that can cross the BBB: alectinib, carboplatin, cobimetinib, danazol, erlotinib, gefitinib, melphalan, panobinostat, ponatinib, rucaparib and vandetanib.

To prioritize the best candidates for DR-TLE, we thoroughly explored their safety profile and the information derived from the previous use of these drugs in epilepsy. For instance, there are clinical case reports and studies showing patients who developed seizures after being treated with melphalan for myeloma ^73^, alectinib for lung tumors ^74^, or vandetinib or gefitinib for glioma ^75,76^. Moreover, gefitinib showed brain toxicity in a chronic epilepsy drug screening ^77^. In addition, it was reported that carboplatin can induce the development of reversible posterior leukoencephalopathy syndrome, which courses with seizures ^78^ and was not suitable for treating chronic epilepsy in a drug screening ^77^. Cobimetinib does not have anticonvulsant effects by itself because, for the treatment of advanced metastatic melanoma, it must be used in combination with carbamazepine to control seizures ^79^. Therefore, based on the previous publications and their safety profile (Supplementary Table 4), we do not recommend validating alectinib, carboplatin, cobimetinib, gefitinib, melphalan, or vandetanib in DR-TLE.

On the other hand, previous publications suggest that there are drug candidates that might be effective for DR-TLE. For instance, erlotinib is approved for tumoral epilepsy ^80^ and is not a substrate of P-glycoprotein, so most of the drug dose will reach the brain. Panobinostat is approved for the treatment of refractory multiple myeloma. It is a histone deacetylase inhibitor, in the same way as valproic acid, and was proposed as a possible antiseizure therapy ^10,81^ and a therapeutic alternative for neuromuscular disease ^82^. Moreover, this drug is promising for DR-TLE since it could modify gene expression and prevent epileptogenesis ^10,81^. Danazol is approved for fibrocystic breast cancer. In one case study danazol was applied to treat cerebral endometriosis after brain surgery and the patient remained seizure-free with this treatment ^83^. Rucaparib, indicated for recurrent epithelial ovarian cancer, was proposed to treat inflammatory diseases, including neurological disorders ^84^, and was even proposed for visual epilepsy in a drug repurposing study ^85^. Ponatinib is used to treat chronic myeloid leukemia. In an *in-silico* study, ponatinib was suggested for the treatment of functional seizures ^86^, and it also reduced seizure severity in a kainate-induced status epilepticus rat model ^87^.

Most of the ASDs are taken in polytherapy for DRE ^88^. Therefore, we studied the ability of the selected drug candidates to inhibit the main members of the CYP450 superfamily (CYP1A2, CYP2C9, CYP3C19, CYP2D6, and CYP3A4). These drugs inhibit many of these enzymes, so they may cause interactions when used in combination with other ASDs. However, it would be safe to combine them with drugs that are not metabolized by the liver, such as levetiracetam ^89^. Another factor we considered was whether our drug candidates are substrates of P-glycoprotein that pumps drugs out of the brain ^90,91^. We found that panabinostat, danazol, ponatinib, and rucaparib are substrates, which may require dose adjustment to achieve the therapeutic dose in the brain. Although these drug candidates may be promising treatment for DR-TLE, they are all part of pharmacological chemotherapies that can cause a wide range of very common ADRs. This may be due to the databases used, such as Pandrugs2, which prioritize anti-tumor drugs ^46^. However, for treating epilepsy, the therapeutic dose may be lower than that required to treat tumors, thus reducing the drugs’ toxicity. Further studies will be needed to determine the adequate dose to treat DR-TLE, but given the severity of DR-TLE, if these drugs decrease the frequency of seizures, the benefits may outweigh the risks.

There is only one other publication that performed drug repurposing based on transcriptomics in epilepsy ^34^. We observed that 67 drugs proposed were common to both studies, such as metformin ^92^ and nifedipine, which were validated in a zebrafish model ^34^. We did not have the chance to validate the results in an animal model which is the main limitation of this study. However, we used 6 different repurposing databases, we studied their ability to cross the BBB, as well as their safety profile, and we relied on a larger cohort than the previous publication (N=46 (our study) vs N=6 ^34^).

In conclusion, we provide insights into the changes in gene expression associated with DR-TLE. This study proposes promising potential drug candidates to treat DR-TLE: danazol, rucaparib, erlotinib, ponatinib, and panobinostat. Nevertheless, further research is needed to validate these results in a preclinical model.

## SUPPLEMENTARY TABLE

**Supplementary Table 1.** Differentially expressed genes (DEGs) associated with drug-resistant temporal lobe epilepsy in the hippocampus sorted by their logarithm of fold change. Genes under-expressed in drug-resistant temporal lobe epilepsy patients with respect to controls have a negative value of Log2FC and are shown in red. Genes over-expressed in drug-resistant temporal lobe epilepsy patients with respect to controls have a positive value of Log2FC are shown in green. Abbreviations: Log2FC: logarithm of fold change.

**Supplementary Table 2.** Definition of the main clusters created by Cytoscape. A) Clusters of proteins encoded by upregulated genes in drug-resistant temporal lobe epilepsy patients with respect to controls. B) Clusters of proteins encoded by downregulated genes in drug-resistant temporal lobe epilepsy patients with respect to controls.

**Supplementary Table 3.** Detailed information about the repurposed drugs obtained with the different databases. A) Drugs proposed by each database based on upregulated genes in drug-resistant temporal lobe epilepsy patients with respect to controls. B) Selection of the drugs proposed by at least 2 different databases based on upregulated genes in drug-resistant temporal lobe epilepsy patients with respect to controls. C) Characteristic of the drugs proposed by at least 2 different databases based on upregulated genes in drug-resistant temporal lobe epilepsy patients with respect to controls. D) Drugs proposed by each database based on downregulated genes in drug-resistant temporal lobe epilepsy patients with respect to controls. E) Selection of the drugs proposed by at least 2 different databases based on downregulated genes in drug-resistant temporal lobe epilepsy patients with respect to controls. F) Characteristics of the drugs proposed by two different databases using downregulated genes in drug-resistant temporal lobe epilepsy patients with respect to controls. We tested drugs’ ability to cross the BBB, whether they were substrates of PgP or PAINS and we showed their SMILES code. Abbreviations: BBB: blood-brain barrier, CMap: Connectivity Map; DGIdb: Drug Gene Interaction Database; PgP: P-glycoprotein, PAINS: Pan-Assay Interference compounds; SMILES: Simplified Molecular Input Line Entry System. Drugs that can cross the blood-brain barrier were highlighted in pink.

**Supplementary Table 4:** Detailed information of the pharmacodynamics, pharmacokinetics and toxicological characteristics of drug candidates. Only very common drug reactions, affecting more than 10% of the population are shown in this table. Promising drug candidates for drug-resistant temporal lobe epilepsy, based on previous publications and safety were shown in green.

Abbreviations: BBB: blood-brain barrier, CYP: cytochrome P 450; GI: gastrointestinal; P-gp: P-glycoprotein.

**Supplementary Figure 1. A)** Enrichment analysis of the downregulated with a logarithm of fold change in drug-resistant temporal lobe epilepsy patients with respect to controls lower than 0.5 using g:Profiler. **B)** Enrichment analysis of the upregulated with a logarithm of fold change in drug-resistant temporal lobe epilepsy patients with respect to controls higher than 0.5 using G-profiler. **C**) Enrichment analysis performed with FUMA GWAS analyzing functions of the DEGs in the hippocampus according to the immunological signature libraries. Red bars represent the proportion of overlapping genes in the gene set. Blue bars show the enrichment p-value, represented as the logarithm of the FDR adjusted p-value. Abbreviation: FDR, false discovery rate.

**Supplementary Figure 2.** Venn diagrams of the comparison and matching of repurposed drugs among different databases. A) Upregulated genes B) Downregulated genes in drug-resistant temporal lobe epilepsy patients with respect to controls. Abbreviations: CMap: Connectivity Map; DGIdb: Drug Gene Interaction Database.

## DATA AVAILABILITY

All data produced in the present study are available upon reasonable request to the authors.

## ACKNOWLEDGMENTS

We are particularly grateful for the generous contribution of the patients and the different Spanish Biobanks: “Biobanco del Hospital Universitario Puerta de Hierro” (HUPHM), “Biobanco del Instituto de Investigaciones Biomédicas August Pi i Sunyer” (IDIBAPS), and “Biobanco de Navarrabiomed” (Navarrabiomed) and “Biobanco en Red de la Región de Murcia” (BIOBANC-MUR). BIOBANC-MUR (Collaboration of Biobank Network of the Region of Murcia) is registered on the Registro Nacional de Biobancos with registration number B.0000859. BIOBANC-MUR is supported by the “Instituto de Salud Carlos III (proyecto PT20/00109), by “Instituto Murciano de Investigación Biosanitaria Virgen de la Arrixaca, IMIB” and by “Consejeria de Salud de la Comunidad Autónoma de la Región de Murcia”.

We would like to thank the following scientists for their help with this study: Dr. Javier Fraga, Dr. Juan Cruz Cigudosa and Gemma Benito. We would like to thank Dr. Gillian Moody for her valuable comments on this manuscript.

## ETHICAL DECLARATION

The protocol and the Informed Consent Form were approved by the Independent Clinical Research Ethics Committee of the Hospital Universitario de La Princesa. The study followed the STROBE guidelines and the Revised Declaration of Helsinki.

## PATIENT CONSENT STATEMENT

All the recruited patients agreed with the protocol and signed the informed consent approved by the Independent Clinical Research Ethics Committee of Hospital Universitario de La Princesa.

