## Supplementary figures and images for "“Transcriptomic Profiling Unveils Novel Therapeutic Options for Drug-Resistant Temporal Lobe Epilepsy”"

### Supplementary Figure 1

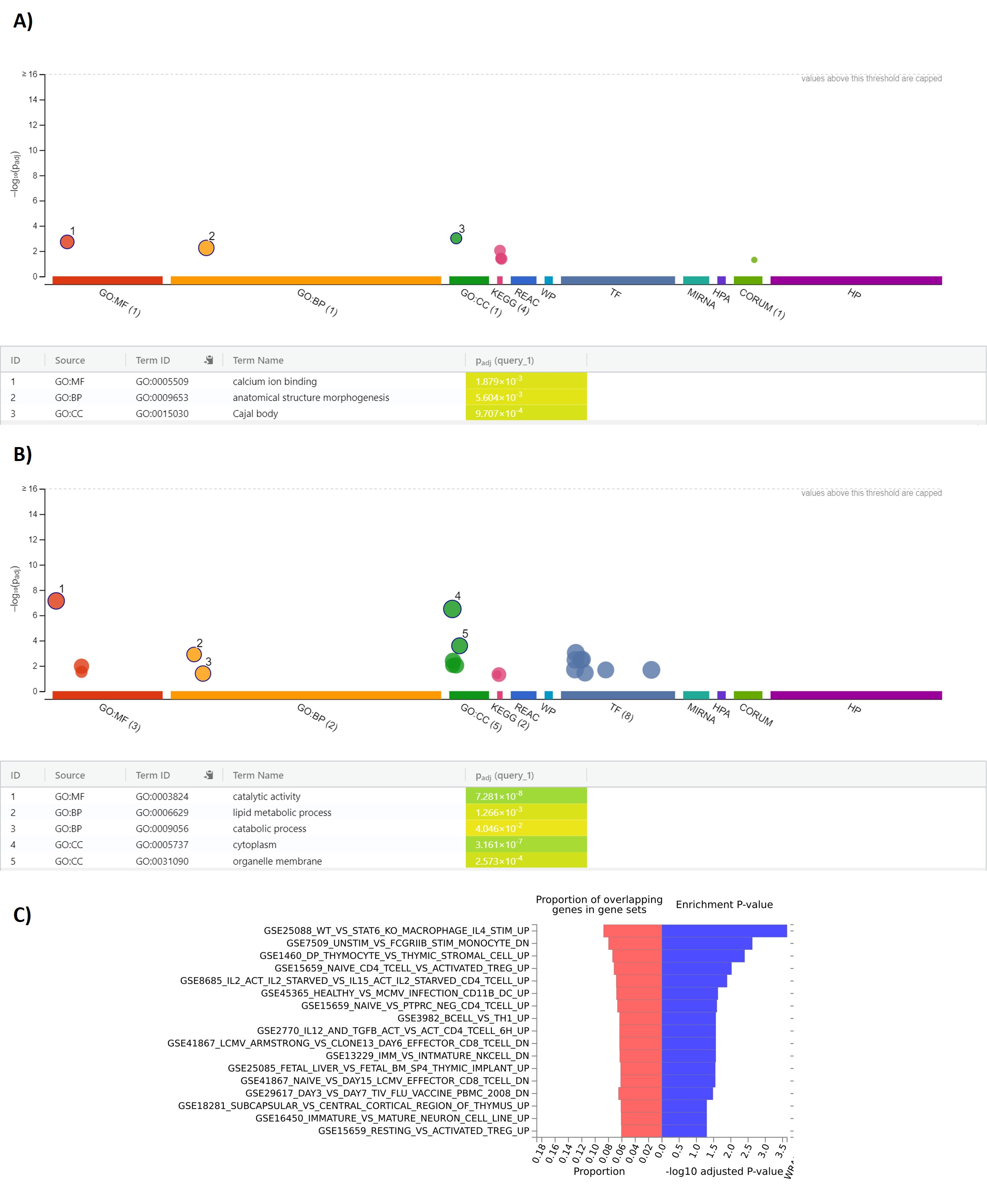

### Supplementary Figure 2

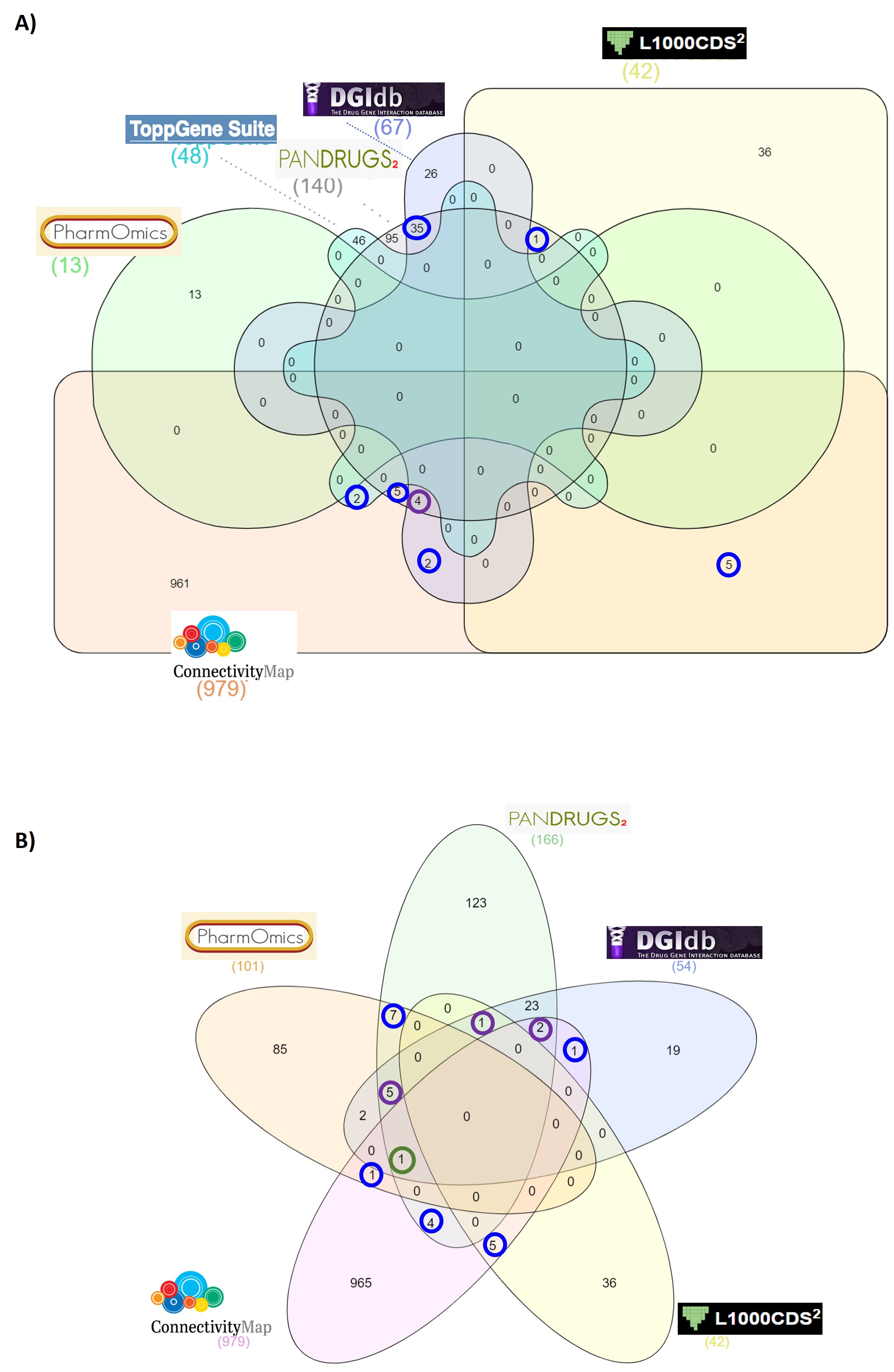
